# Are we testing for triple elimination? A review of national guidelines on preventing mother-to-child transmission of HIV, syphilis, and hepatitis B in the African region

**DOI:** 10.1101/2024.10.29.24316372

**Authors:** Kellie List, Aliza Monroe-Wise, Margaret Banks, Magdalena Barr-DiChiara, Agnes Chetty, Morkor Newman-Owiredu, Olufunmilayo Lesi, Cheryl Case Johnson, Alison L. Drake

## Abstract

**Introduction:** World Health Organization (WHO) updated guidelines for maternal HIV, syphilis and hepatitis B virus (HBV) testing for pregnant women, and advised on using dual HIV/syphilis rapid diagnostic tests (RDTs). We reviewed national testing guidelines to assess political support in alignment with WHO guidance and triple elimination of mother-to-child transmission (MTCT) priorities.

**Methods:** We reviewed national HIV-related guidelines from 46 African countries after 2010, including guidelines on prevention MTCT, HIV testing/self-testing, HIV antiretroviral treatment, and/or HIV pre-exposure prophylaxis. Data was extracted data on recommendations for maternal testing for HIV (frequency and timing), syphilis (any test/test type); and HBV. Testing recommendations for all three infections in the most recent country guidelines were compared to WHO guidelines.

**Results:** Testing policies on HIV, syphilis, or HBV during pregnancy and/or postpartum from 38 (83%) countries were identified; 18 (47%) had policies updated after 2019 and 11 (29%) were high HIV burden. All 38 countries with HIV testing guidelines addressed maternal HIV retesting, while 68% had guidelines recommending syphilis testing (46% of which recommended the dual RDT), and 58% had HBV testing guidelines. Maternal HIV retesting was more frequent than recommended by WHO in 2019 in 9/11 (82%) and 7/27 (26%) of high and low HIV burden countries, respectively. Half of countries had triple elimination of MTCT focused policies, and 10 (26%) had maternal HIV retesting as well as syphilis and/or HBV testing guidance.

**Conclusions:** Half of countries had guidelines to test for all three infections; policy evaluations to measure implementation and impact are needed.

## INTRODUCTION

Triple elimination of mother-to-child transmission (EMTCT) of HIV, syphilis, and hepatitis B virus (HBV) is a critical priority due to the morbidity and mortality associated with mother-to-child transmission (MTCT) with these infections. Approximately 1.3 million women living with HIV become pregnant every year, 900,000 pregnant women are infected with syphilis globally, and 83% of new HBV infections are in Africa or South-east Asia [1]. In 2019, the World Health Organization (WHO) guidelines highlighted maternal testing for HIV, syphilis and HBV, including the use of the dual HIV/syphilis rapid tests [2]. Despite this, national policies and implementation vary widely, with competing priorities and resource limitations acting as barriers to progress.

The prevention of MTCT has seen the most progress in HIV, largely due to dedicated investments which had resulted in widespread HIV testing in antenatal care. As a result, EMTCT efforts will need to build upon the existing international and national policy and funding infrastructure for HIV testing [3].

In contrast, coverage of syphilis and HBV testing in pregnant people lags behind that of HIV. Despite recommendations to test all pregnant people for all three infections, disparities in funding and advocacy have contributed to lower syphilis and HBV testing coverage. In response to this gap, WHO recommends dual HIV/syphilis rapid diagnostic tests (RDTs) and HBV testing for all pregnant people at least once in pregnancy. Dual testing, requiring only one test for both HIV and syphilis infections, can be cost-saving or cost-effective, and capitalize on the high HIV testing coverage to deliver more syphilis testing and treatment to pregnant women [2,4]. Fewer resources and testing options are available for HBV, compared to HIV, making it challenging to progress toward EMTCT goals.

For countries with a high HIV burden, strategies such as catch-up testing for those who miss or delay visits, as well as retesting in the third trimester, are essential components of EMTCT efforts [2]. PrEP is also safe and should be encouraged, given the increased risk of HIV acquisition during these periods [5]. Moreover, partner testing and self-testing approaches have proven effective in reaching male partners of pregnant women [6-7]. However, our prior review of maternal HIV testing guidelines revealed that many countries recommend retesting at suboptimal intervals and frequencies that do not align with WHO guidelines [8]. A modeling study further suggested that retesting more frequently than necessary or at inappropriate times can lead to higher costs with limited impact on HIV transmission prevention [9].

To advance the integration and scale-up of testing for all three infections in alignment with WHO guidance, it is important to assess whether national guidelines support EMTCT goals. This review of guidelines from African countries on maternal testing for HIV, syphilis, and HBV will identify gaps and inform policy planning to enhance implementation efforts and promote EMTCT.

## METHODS

HIV guidelines from 46 countries in Africa were reviewed and compared with the 2019 WHO guidelines. Available guidelines published in 2019 or after on prevention of mother-to-child HIV transmission (PMTCT), HIV testing, HIV self-testing, HIV antiretroviral treatment (ART), and/or HIV pre-exposure prophylaxis (PrEP) were included in the review.

Country guidelines were provided by WHO. If countries had multiple guideline documents, we used the most recently published version, or the guideline with the most complete information for each piece of information extracted. We electronically searched country guidelines for terms related to the following topics: HIV testing and retesting, postpartum, pregnancy, syphilis testing, and HBV testing, pregnancy, labour/delivery, postpartum, or lactating/breastfeeding.

No country guidelines were excluded from the current review based on language. Non-English languages included Portuguese, French, and Spanish. All non-English guidelines were reviewed by the first reviewer (MB) using Google translated versions of the guidelines. Reviewers from WHO, fluent in each language, were consulted to interpret translations when necessary.

Primary guideline reviews were conducted by two individuals (MB and KL). Primary reviewers extracted data into a standardized data collection form and switched roles to serve as secondary reviewers to verify abstraction from the other reviewer; discrepancies were resolved by a third reviewer (AD). Data from policies was abstracted on: HIV testing at initial antenatal care (ANC) visit; maternal and partner HIV self-testing; partner HIV and syphilis testing; HIV retesting during pregnancy, labour/delivery, and/or postpartum; syphilis testing during pregnancy and/or labour/delivery; syphilis test type (rapid diagnostic tests [RDTs] or lab-based testing); confirmatory syphilis tests; HBV testing during pregnancy and/or labour/delivery, HBV test type, and HBV household testing.

### HIV retesting

Countries were classified as having high (prevalence >5%) or low (prevalence ≤5%) HIV burden as previously described based on the most recent 2022 UNAIDS estimates available for women aged 15-49 years, or for adults aged 15-49 years if not available. Maternal HIV retesting was defined as testing after the first ANC visit (third trimester, at labour/delivery, and/or postpartum). Countries with general population policies for retesting (not specific to pregnancy or postpartum) were classified as not having guidance on maternal HIV retesting.

We also classified countries based on the frequency of HIV retesting recommended by WHO. For high HIV burden countries, HIV retesting was classified as 1) less frequent than the WHO guidelines if they do not recommend retesting at least once during late pregnancy or at labor and delivery and no retesting in the postpartum period, 2) as frequent as the WHO guidelines if they recommended retesting once during late pregnancy (and/or at labor and delivery) and/or 1-2 times in the postpartum period, 3) more frequent than the guidelines if they recommended retesting more than once during late pregnancy (and/or at labor and delivery) and/or more than 2 times in the postpartum period.

For low HIV burden countries, maternal HIV retesting is not universally recommended by WHO. In these settings it is advised that HIV-negative women be retested in the third trimester if they are in serodiscordant relationships with partners on ART with unsuppressed viral load, or have ongoing risk; and during the postpartum period for women from these groups or who are from a key population. For low HIV burden settings, countries are classified as 1) less frequent than guidelines if they do not recommend retesting for these special groups of women, 2) as frequent if they recommend one retest during late pregnancy, with or without an additional postpartum retest, and 3) more frequent for countries who recommend >1 retest during late pregnancy and/or >1 retest postpartum for these special groups of women or for all women.

### Syphilis testing

Syphilis testing was classified as laboratory based, individual RDT, or dual RDT. The laboratory test types included venereal disease research laboratory test (VDRL), rapid plasma reagin (RPR), or treponema pallidum hemagglutination assay (TPHA). RDTs included both the individual syphilis RDT or the dual HIV/syphilis RDT. Confirmatory testing for syphilis was defined as any test (laboratory or RDT) to confirm a positive test result. We extracted data on whether syphilis testing was recommended at specific time points during pregnancy and postpartum, and if male partner testing was included.

### HBV testing

We classified guidelines based on populations recommended for HBV testing as follows: general population, pregnant women, people living with HIV (PLHIV), pregnant women with HIV, and key populations. We captured data on HBV test type, and HBV household testing, which is inclusive of partner testing recommendations.

## RESULTS

### Overview of country policies

Of 46 countries in the review, 12 (26%) were from high HIV burden settings. Overall, 38 (83%) countries had integrated HIV testing guidelines covering HIV, syphilis, and/or HBV during pregnancy and/or postpartum periods (Table 1); 18 (47%) of these had guidelines updated after 2019. Three of eight countries (3 low HIV burden) with no testing guidelines for any of the three infections were updated after 2019 while five countries (1 high HIV burden, 4 low HIV burden) had guidelines published prior to or during 2019.

**Table 1.**
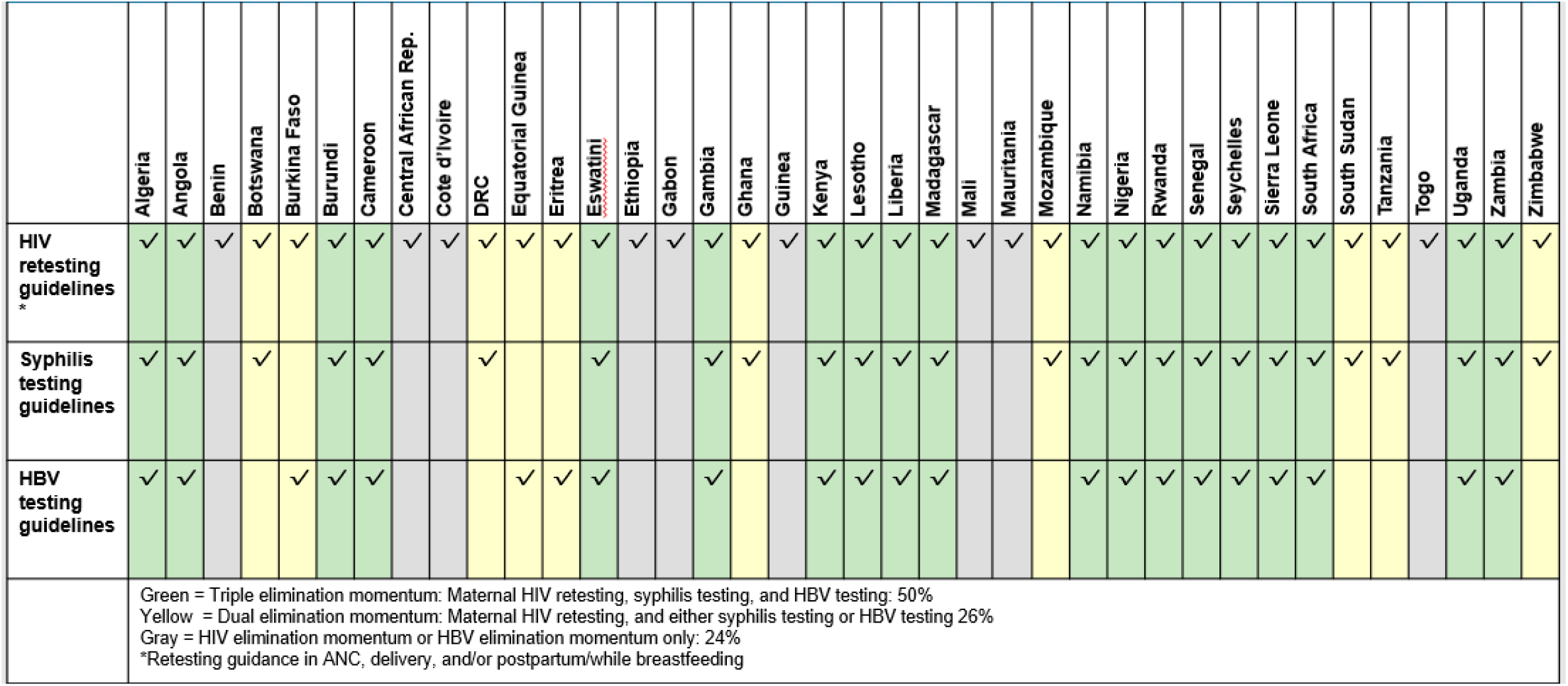
Maternal testing guidelines supporting triple EMTCT, by country (n=38)

### HIV retesting

All 38 countries with policies identified had maternal HIV retesting guidelines. Overall, 18 (47%) had retesting guidelines that aligned with WHO guidance, while 16 (42%) had guidelines that recommended more frequent testing, and 3 (8%) were less frequent. Among 11 high HIV burden countries, no countries were fully aligned with WHO guidelines, while 2 (18%) were less frequent and 9 (82%) were more frequent. In contrast, among 27 low HIV burden countries, 19 (70%) were as frequent as WHO guidance, while 1 (3%) was less frequent and 7 (26%) were more frequent. Among 18 (6 high and 12 low HIV burden) countries with guidelines updated after 2019, 8 (44%) were more frequent (5 high and 3 low HIV burden) while 9 (50%) were as frequent (9 low HIV burden) and 6% (1 high HIV burden) were less frequent than WHO guidelines. Of 38 countries with any testing guidelines, 58% had guidelines on maternal HIV self-testing (HIVST), 74% had partner HIV testing guidelines, and 61% had guidelines on PrEP in pregnancy.

### Syphilis testing

Among 38 countries with available HIV guidelines, 26 (68%) also had guidelines for pregnant women to be tested for syphilis: 6 (23%) identified lab-based tests, 12 (46%) recommended dual RDT (3 of 12 [25%] which recommend lab-based confirmatory syphilis testing for diagnosis for women with reactive RDT results), and 7 (27%) did not specify the test type (Figure 1). Among the 12 countries who recommend the dual RDT, all 12 recommend it as the first ANC test (46% of all countries with syphilis test recommendations). Among the 18 countries with guidelines updated after 2019, 15 (83%) recommend syphilis testing in pregnancy, 11 of which (61%) recommended the dual RDT. In addition, 35% of countries with syphilis testing recommended syphilis partner testing and 42% recommended syphilis retesting in pregnancy.

**Figure 1.**
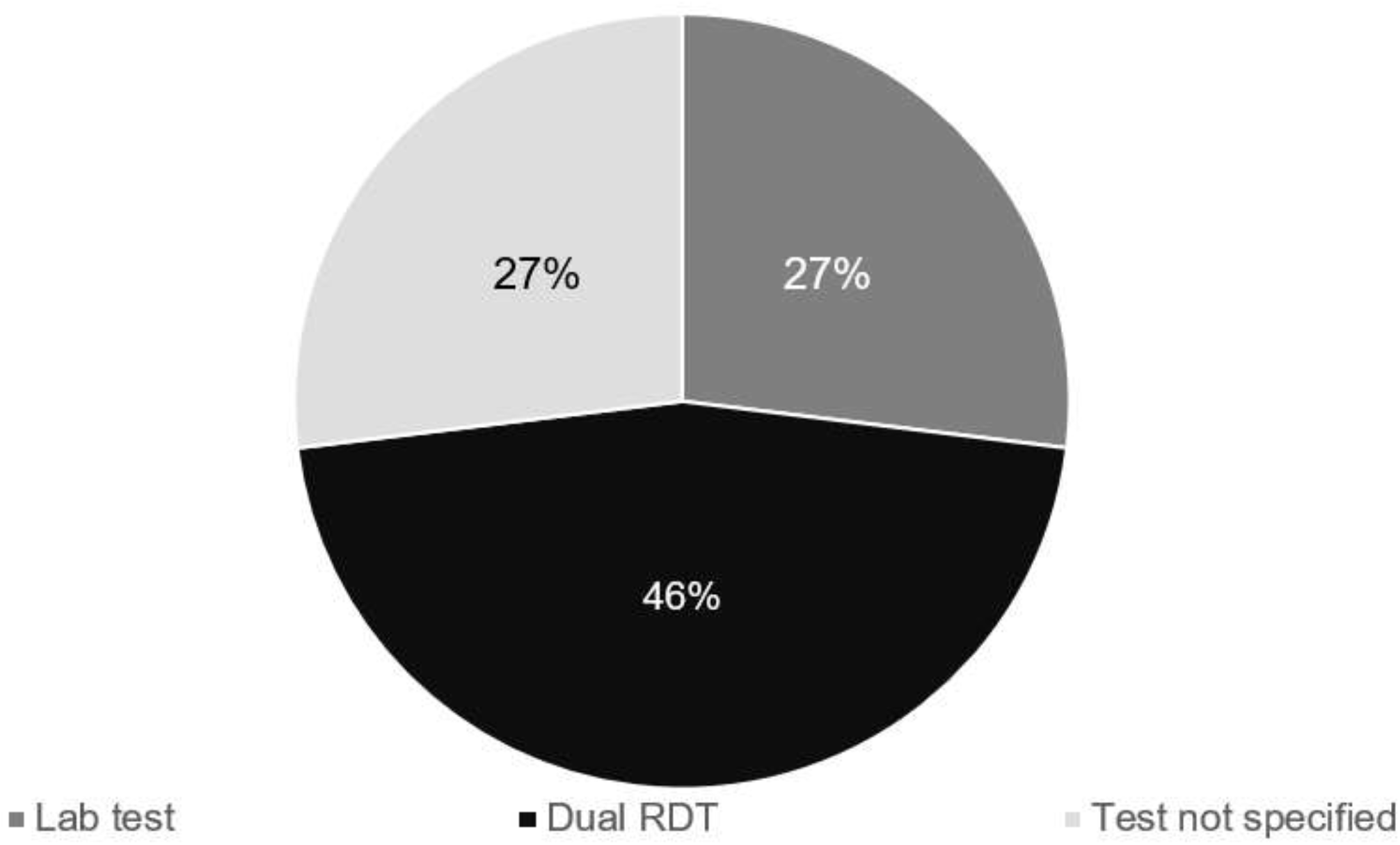
Syphilis test type in pregnancy (n=26)* *12 of 38 countries (32%) with any guidelines were excluded as they did not have syphilis testing guidelines.

### HBV testing

Twenty-two (58%) of 38 countries with any guidelines recommended maternal HBV testing, 13 (59%) of which specified using HBsAg specifically, while the other countries did not indicate test type. In addition, three countries (14%) recommended testing of household members.

### Progress towards testing for triple EMTCT

Among 38 countries with any testing guidance, 19 (50%) have maternal HIV retesting, syphilis testing, and HBV testing guidelines; Ten additional countries (26%) have maternal HIV retesting, and either syphilis testing or HBV testing guidelines; 9 (24%) only had maternal HIV retesting guidance (Table 1).

## DISCUSSION

Our review of national guidelines across sub-Saharan Africa indicates that while significant progress has been made toward EMTCT of HIV, syphilis, and HBV, gaps remain in policy adoption and implementation. The high coverage of HIV testing and maternal retesting reflects strong political and financial commitments, built on decades of international support for HIV prevention. This progress has not been mirrored in syphilis and HBV testing, however, which lag behind.

A key finding is that while 83% of countries with policies reviewed had PMTCT-related testing guidance for HIV, syphilis, and/or HBV, a little more than half of the county policies reviewed had guidance for HIV retesting, syphilis or HBV during pregnancy, signaling uneven efforts toward triple elimination. The lower adoption of syphilis and HBV testing guidelines highlights disparities in funding streams, advocacy, and the prioritization of non-HIV infections within maternal care frameworks.

HIV retesting practices appeared to vary widely, with some countries retesting too frequently and at suboptimal time points, leading to inefficient use of resources. Our review echoes modeling studies that suggest increasing the frequency of HIV retesting beyond WHO recommendations does not result in meaningful public health impact [4, 9]. Conversely, in 18% of high HIV burden countries, HIV retesting occurs too infrequently. These discrepancies suggest greater efforts to align national policies and allocation of resources are needed. Redirecting resources from excessive HIV retesting to increase syphilis and HBV testing could be more impactful and further accelerate progress toward EMTCT.

Despite the well-documented morbidity and mortality associated with congenital syphilis, maternal syphilis testing remains significantly lower than HIV testing [10]. Integrated HIV and syphilis testing, particularly with rapid or self-tests which enable same day diagnosis and treatment, have been proposed as a way to overcome existing testing barriers (e.g. limited access to lab-settings and long wait times). Dual tests are cost-effective and efficient [11-13], yet only 62% of countries have adopted them within antenatal care [15]. This low uptake, coupled with the fact only 42% of countries updated their syphilis testing policies following the 2019 WHO guidelines, indicates missed opportunities to expand dual testing persist. With three WHO prequalified dual tests, some under $1.00, integration into national policies must be prioritized to address stagnant and rising congenital syphilis rates [16].

HBV testing, key to triple elimination, has also seen limited uptake [17-18]. Only a little more than half (58%) of countries with HIV guidelines had specific guidance HBV testing in pregnancy; even fewer provided guidance on follow-up tests (e.g. HBsAg or HBV DNA) to guide antiviral prophylaxis. Given the high HBV burden in sub-Saharan Africa, the lack of testing guidelines represents a critical gap for the future of EMTCT efforts in the region. With the 2024 WHO update on HBV testing and treatment [19], it will be important to review policy changes and assess their impact on HBV transmission rates.

Our review highlights both progress and gaps in testing policies for HIV, syphilis, and HBV. Testing serves as the entry point for prevention and treatment, making it crucial to EMTCT. While triple elimination is a global priority, our study was limited by its focus on sub-Saharan Africa countries with available testing-related guidelines. It also did not include sexually transmitted infections (STIs) more broadly or HBV-specific guidelines beyond those related to HIV. Nonetheless, the findings underscore the importance of integrating testing policies across all three infections to optimize maternal and infant health outcomes and contribute to triple elimination.

To move toward EMTCT of HIV, syphilis, and HBV, countries must not only update their testing guidelines to align with WHO recommendations, but also ensure effective implementation. Optimizing and sustaining existing, as well as targeted expansion, of HIV testing during pregnancy remains critical for elimination goals. In parallel, addressing the gaps in syphilis and HBV testing requires greater focus, advocacy, policy updates and dedicated funding. By strengthening and integrating testing frameworks across all three infections, we can reduce infant morbidity and mortality from these preventable infections. Efforts are now needed to achieve the WHO global health sector strategy to end AIDS and ongoing STIs and viral hepatitis epidemics by 2030 [20].

## Data Availability

All data produced in the present study are available upon reasonable request to the authors.

